# Expanding cholera serosurveillance to vaccinated populations

**DOI:** 10.1101/2025.03.09.25323598

**Authors:** Forrest K. Jones, Taufiqur R. Bhuiyan, Damien M. Slater, Ralph Ternier, Kian Robert Hutt Vater, Ashraful I. Khan, Fahima Chowdhury, Kennia Visieres, Rajib Biswas, Mohammad Kamruzzaman, Edward T. Ryan, Stephen B. Calderwood, Regina C. LaRocque, Richelle C. Charles, Daniel T. Leung, Justin Lessler, Louise C. Ivers, Firdausi Qadri, Jason B. Harris, Andrew S. Azman

**Affiliations:** Department of Epidemiology, Johns Hopkins Bloomberg School of Public Health, Baltimore, USA; Infectious Diseases Division, International Centre for Diarrhoeal Disease Research, Bangladesh (icddr,b), Dhaka, Bangladesh; Division of Infectious Diseases, Massachusetts General Hospital, Boston, USA; Zanmi Lasante, Port au Prince, Haiti; Department of Medicine, Harvard Medical School, Boston, Massachusetts, USA; Department of Immunology and Infectious Diseases, Harvard T.H. Chan School of Public Health, Boston, Massachusetts, USA; Division of Infectious Diseases, University of Utah School of Medicine, Salt Lake City, Utah, USA; Division of Microbiology and Immunology, University of Utah School of Medicine, Salt Lake City, Utah, USA; Department of Epidemiology, University of North Carolina Gillings School of Global Public Health, Chapel Hill, NC, USA; Carolina Population Center, The University of North Carolina at Chapel Hill, Chapel Hill, NC, USA; Center for Global Health, Massachusetts General Hospital, Boston, USA; Department of Global Health and Social Medicine, Harvard Medical School, Boston, USA; Harvard Global Health Institute, Cambridge, USA; Department of Pediatrics, Harvard Medical School, Boston, Massachusetts, USA; Centre for Emerging Viral Diseases, Geneva University Hospitals, Geneva, Switzerland; Division of Tropical and Humanitarian Medicine, Geneva University Hospitals, Geneva, Switzerland

## Abstract

Mass oral cholera vaccination campaigns targeted at subnational areas with high incidence are central to global cholera elimination efforts. Serological surveillance offers a complementary approach to address gaps in clinical surveillance in these regions. However, similar immune responses from vaccination and infection can lead to overestimates of incidence of infection. To address this, we analyzed antibody dynamics in infected and vaccinated individuals to refine seroincidence estimation strategies for partially vaccinated populations. We tested 757 longitudinal serum samples from confirmed *Vibrio cholerae* O1 cases and uninfected contacts in Bangladesh as well as vaccinees from Bangladesh and Haiti, using a multiplex bead assay to measure IgG, IgM, and IgA binding to five cholera-specific antigens. Infection elicited stronger and broader antibody responses than vaccination, with rises in cholera toxin B-subunit (CTB) and toxin-coregulated pilus A (TcpA) antibodies uniquely associated with infection. Previously proposed random forest models frequently misclassified vaccinated individuals as recently infected (over 20% at some time points) during the first four months post-vaccination. To address this, we developed new random forest models incorporating vaccinee data, which kept false positive rates among vaccinated (1%) and unvaccinated (4%) individuals low without a significant loss in sensitivity. Simulated serosurveys demonstrated that unbiased seroincidence estimates could be achieved within 21 days of vaccination campaigns by ascertaining vaccination status of participants or applying updated models. These approaches to overcome biases in serological surveillance enable reliable seroincidence estimation even in areas with recent vaccination campaigns enhancing the utility of serological surveillance as an epidemiologic tool in moderate-to-high cholera incidence settings.

**Significance statement:** Serological surveillance can improve how we monitor cholera in high burden areas where clinical surveillance is limited. However, vaccination can produce immune responses similar to infection, leading to overestimates in seroincidence. This study extends seroincidence estimation techniques using machine learning models to partially vaccinated populations. We analyzed antibody dynamics from vaccinated and infected individuals to develop methods that reduce misclassification of vaccinated individuals as recently infected. These methods enable reliable seroincidence estimates in areas with recent vaccination campaigns, providing a step toward better epidemiologic monitoring in the context of global cholera control initiatives.

## Introduction

Cholera remains a global public health threat, with an estimated 95,000 deaths per year (1). The Global Taskforce for Cholera Control’s End Cholera 2030 Roadmap is based on highly focused disease prevention and control in subnational ‘cholera hotspots’ (2). Killed oral cholera vaccines (OCV) are one recommended component of cholera prevention in hotspots as they reduce severe cholera symptoms and limit transmission (3, 4). Clinical surveillance for *V. cholerae* infection is often limited in highly affected communities, with most infections being missed and many suspected cases of diarrheal illness being misattributed as cholera (5, 6). Serological surveillance (i.e. serosurveillance) has been proposed as a complementary approach to measure cholera transmission and burden (7). However, it is unclear whether current lab and analytic methods for serosurveillance are appropriate in vaccinated populations, where immune responses to vaccines could be misattributed to natural infections.

Similarities between the human antibody response to *V. cholerae* infection and OCV have been previously assessed (8). The two most commonly used bivalent OCVs in cholera endemic regions (Shanchol and Euvichol) contain five inactivated strains of *V. cholerae* (both the O1 Inaba and Ogawa serotypes as well as the now extremely rare O139 serogroups) and no component of the cholera toxin (9). The vaccine is administered to individuals over the age of 1 year in two doses spaced at least two weeks apart, though one-dose use has become increasingly common due to the global shortage of vaccines (10). Mass campaigns are generally conducted in two rounds, with each round lasting only a few days to a week (11). Vaccination boosts many of the same serological markers that have been proposed to identify previously infected individuals (7, 12). Therefore, seroincidence estimates from vaccinated populations could overestimate the true infection incidence by misclassifying vaccinated individuals as recently infected, potentially threatening the validity of this approach in cholera endemic areas globally.

Several strategies related to serosurvey design, laboratory protocols, and statistical analyses might mitigate the overestimation of seroincidence in vaccinated populations. In this study, we provide new data on antibody kinetics following vaccination and *V. cholerae* O1 infection to quantify the extent of misclassification of vaccinated individuals as recently infected. We propose and evaluate approaches to enhance seroincidence estimation, including optimizing the timing of serosurveys, developing new machine learning models, and applying targeted adjustment methods. As the use of both serosurveillance and vaccination expands, these findings are crucial for improving the accuracy of serologic data interpretation and supporting cholera control programs. Furthermore, this work provides a framework for evaluating and adapting serosurveillance to partially vaccinated populations for other vaccine-preventable diseases that elicit short-lived systemic IgG antibody responses.

## Results

We analyzed data from 236 samples collected from 43 Bangladeshi vaccine recipients and 212 samples from 36 Haitian vaccine recipients, all of whom received two doses of the Shanchol vaccine, spaced 14 days apart (Table 1). Among the Bangladeshi participants, nearly half (47%) were under 10 years old, and 49% were male. In contrast, all Haitian participants were adults (≥18 years old), with the majority (72%) being male.

**Table 1:**
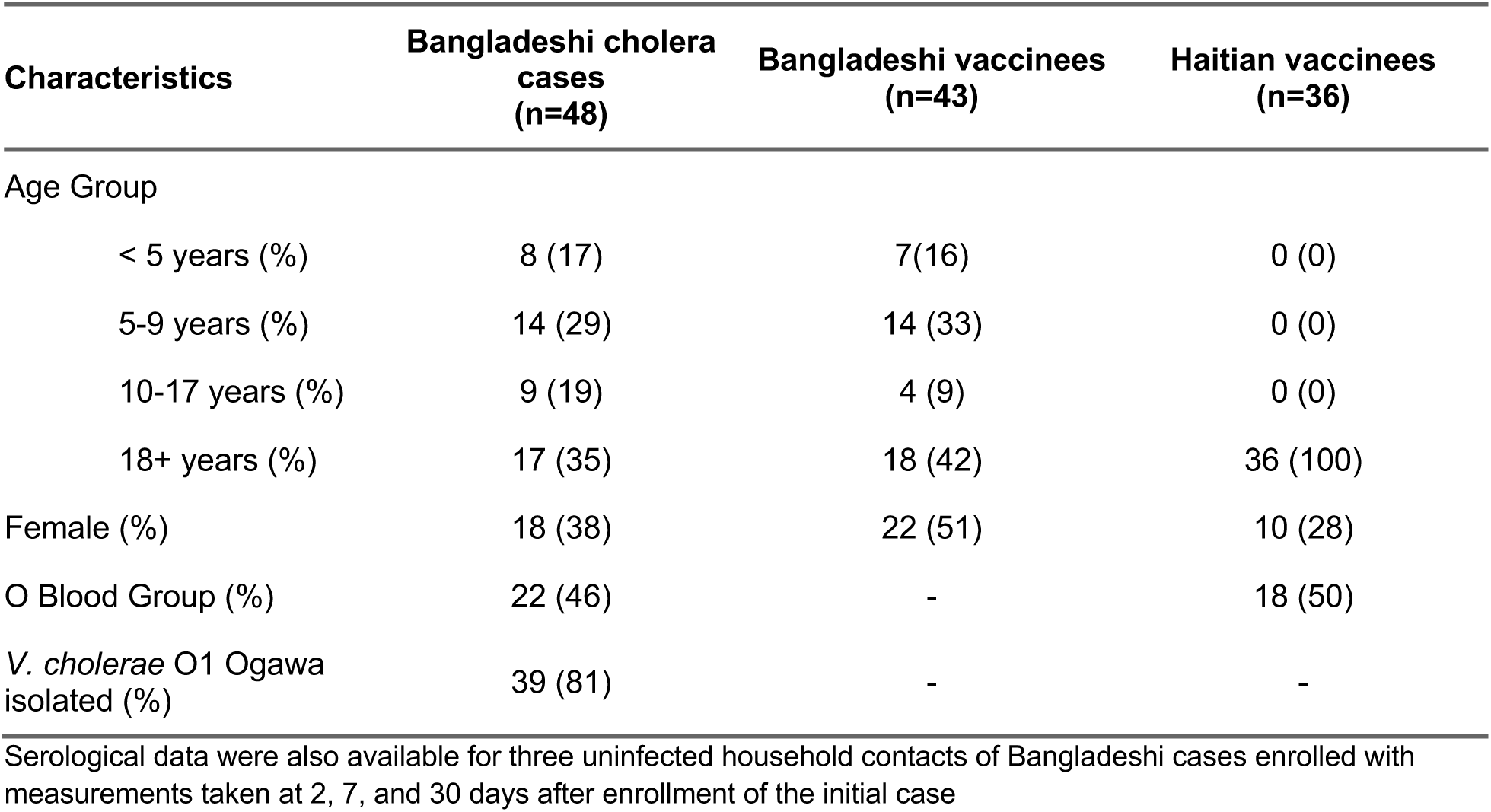
Individual characteristics of culture confirmed cholera patients and vaccinated individuals.

| Characteristics | Bangladeshi cholera cases (n=48) | Bangladeshi vaccinees (n=43) | Haitian vaccinees (n=36) |
| --- | --- | --- | --- |
| Age Group |  |  |  |
| < 5 years (%) | 8 (17) | 7 (16) | 0 (0) |
| 5-9 years (%) | 14 (29) | 14 (33) | 0 (0) |
| 10-17 years (%) | 9 (19) | 4 (9) | 0 (0) |
| 18+ years (%) | 17 (35) | 18 (42) | 36 (100) |
| Female (%) | 18 (38) | 22 (51) | 10 (28) |
| O Blood Group (%) | 22 (46) | - | 18 (50) |
| *V. cholerae* O1 Ogawa isolated (%) | 39 (81) | - | - |

Serological data were also available for three uninfected household contacts of Bangladeshi cases enrolled with measurements taken at 2, 7, and 30 days after enrollment of the initial case

Additionally, we included data from a Bangladeshi cohort of patients with *V. cholerae* infection, consisting of 300 samples from 48 individuals, primarily young (46% under 10 years old) and predominantly male (62%). Most of these individuals (81%) had *Vibrio cholerae* O1 serotype Ogawa isolated from their stool, while the remainder were of the Inaba serotype. We also analyzed nine samples from three uninfected household contacts of patients with cholera.

Baseline samples were collected from all vaccinated individuals on the day of their first dose. Bangladeshi vaccine recipients provided additional samples up to 42 days post-vaccination, while Haitian participants were followed for a longer duration, with samples collected up to 360 days post-vaccination (Table S1). Patients with *V. cholerae* O1 infection were followed for up to 1,080 days, with periodic blood sample collection (2–7 samples per individual). Household contacts each provided three serum samples within the first 30 days of follow-up.

### Immune response to vaccination and infection differ in magnitude and breadth

Bangladeshi patients with naturally occurring *V. cholerae* O1 infections and vaccinees had similar baseline IgG antibody distributions after stratifying by age group (<10 years vs 10+ years, Figure 1, Figure S1). Haitian vaccinees had lower baseline levels of anti-OSP IgG antibodies against both serotypes, Inaba and Ogawa. While both infection and vaccination led to large rises in anti-OSP antibodies against both serotypes, infection led to a more consistent and robust rise against both serotypes than vaccination (Figure 1, Table S2 and S3). IgA and IgG antibodies targeted at the cholera toxin (B subunit, CTB) rose dramatically after infection but not vaccination, with neither exposure type leading to a rise in anti-CTB IgM. Despite the presence of *Vibrio cholerae* O139 in the vaccine, only a minority of vaccinees had more than a 2-fold rise in anti-O139 OSP antibodies across isotypes (18% IgM, 22% IgG, and 42% IgA). Most cases (75% and 67%) but few vaccinees (30% and 18%) generated a greater than 2-fold rise in anti-TcpA IgG and IgA. We used multidimensional scaling to map the dynamic antibody profiles of those recently infected or vaccinated (200 days) and found that they form partially-separated clusters (Figure S2).

**Figure 1:**
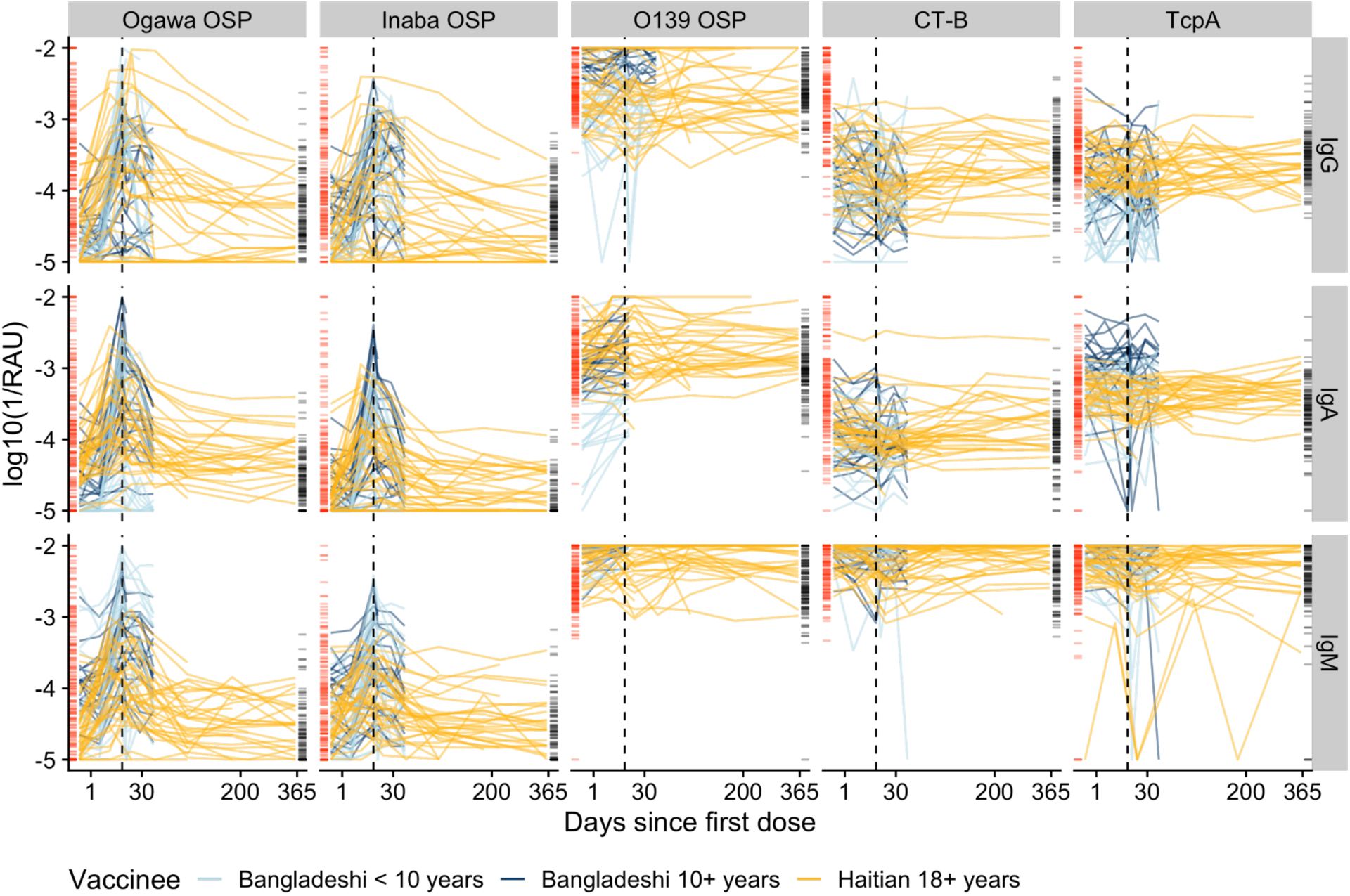
Multiplex bead assay measurements of IgG, IgM, and IgA against OSP, CTB, and TcpA antigens among vaccinated volunteers and comparison with cholera cases. The Y-axis indicates the concentration of antibodies shown as the log (base 10) of the inverse relative antibody unit (RAU). The X-axis, days since the first dose of vaccination, is square-root transformed. Each colored line indicates individual trajectories over time (light blue: Bangladeshi vaccinees <10 years, dark blue: Bangladeshi vaccinees ≥10 years, gold: Haitian vaccinees ≥18 years). Rug plots on the vertical axes show the antibody measurements from the cohort of cholera cases in Bangladesh (red: case measurements inside the 200-day infection window; black: uninfected household contacts and case measurements outside a 200-day infection window). The black dotted line indicates the timing of second dose vaccination, 14 days after the first dose.

### Previous seroincidence models frequently misclassify vaccination as recent infection

We assessed how often vaccinated people were misclassified as seroincident (i.e., predicted to be recently infected based upon a serological response profile) using a previously published seroincidence model. This model, the *Infection-Only* model (Table 2), was trained on three IgG antibody measurements (Ogawa OSP, Inaba OSP, and CTB) from unvaccinated individuals in Bangladesh. We investigated four post-infection time periods (i.e., infection windows) where infections were considered recent: 45, 120, 200 and 300 days. These models were calibrated to classify no more than 5% of people not recently infected as seroincident (i.e., a nominal false positivity rate of 5%). We found that vaccinees were rarely misclassified as seroincident, but it depended on the infection window used and the time since vaccination.

**Table 2.** Seroincidence model descriptions. *Serological data from three uninfected household contacts of Bangladeshi cases were also used

| **Name** | **Training Data Source** | **Markers** | **Classes** |
| --- | --- | --- | --- |
| *Case-Only Model* | Bangladeshi cholera patients* | anti-CTB IgG<br>anti-Ogawa OSP IgG<br>anti-Inaba OSP IgG | Recently infected<br>Not recently infected |
| *Mixed-Cohort Two Class Model* | Bangladeshi cholera patients*<br>Bangladeshi vaccinees<br>Haitian vaccinees | anti-CTB IgG<br>anti-Ogawa OSP IgG<br>anti-Inaba OSP IgG | Recently infected<br>Not recently infected |
| *Mixed-Cohort Three Class Model* | Bangladeshi cholera patients*<br>Bangladeshi vaccinees<br>Haitian vaccinees | anti-CTB IgG<br>anti-Ogawa OSP IgG<br>anti-Inaba OSP IgG | Recently infected<br>Recently vaccinated<br>Neither |

Models using a short infection window (e.g., 45-days) rarely misclassified vaccinees, though increasing the infection window led to longer periods of elevated misclassification (Figure 2). With a 120-day infection window, this period lasted 49 days, peaking at 12% misclassification. The period of elevated misclassification grew longer (111 days and 114 days) with higher levels (24% and 23%) of misclassification when using models with 200- and 300-day infection windows. Misclassification did not differ substantially between the Haitian and Bangladeshi vaccinees (Figure 2). Previously proposed models based on a larger set serological markers and isotypes (IgA and IgM), including anti-TcpA and anti-O139 OSP antibodies, did not perform substantially better than those based on three IgG markers, and for shorter time windows they led to greater misclassification (Figure S4).

**Figure 2:**
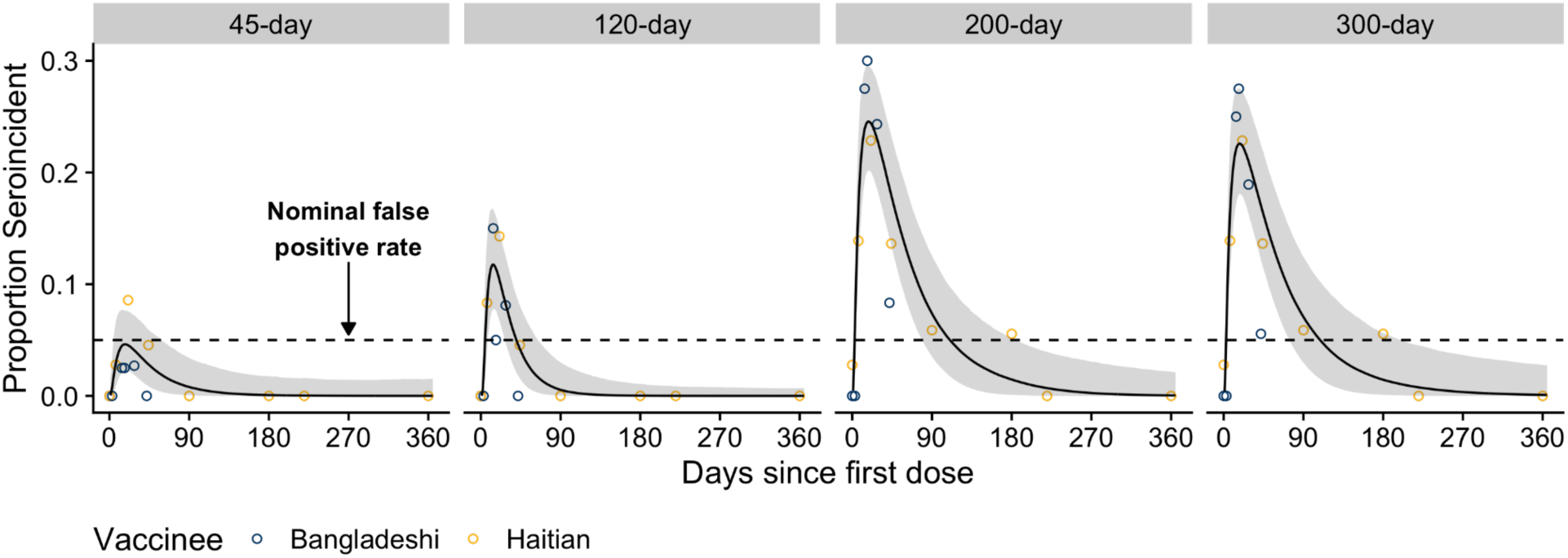
Misclassification of vaccinees as seroincident by previous seroincidence models using 45, 120, 200, and 300 day infection windows. The model used (i.e., the *Infection-Only* Model) to classify vaccinees as seroincident or not was a previously published random forest model trained on anti-CTB, anti-Ogawa OSP, anti-Inaba OSP IgG measurements from Bangladeshi confirmed cases and uninfected household contacts. The proportion of Bangladeshi (dark blue) and Haitian vaccinees (gold) classified as seroincident are shown as dots. The overall proportion seroincident was modeled with a cubic spline (black line and grey ribbon) using data from both cohorts of vaccinees. Black dashed line indicates the nominal false positivity rate of 5%.

### Inclusion of data from vaccinees improved performance of seroincidence models without the need for additional antigen targets

We then evaluated the performance of two new random forest models to identify individuals infected in the last 200 days when trained with additional data from vaccinees. The *Mixed-Cohort Two Class Model* was trained on cases, uninfected household contacts and vaccinees, and classified individuals as being recently infected or not recently infected (Table 2). To distinguish between recently infected, recently vaccinated (<200 days), and neither recently vaccinated/infected, we developed the *Mixed-Cohort Three Class Model*, which we trained on the same data. Seroincidence models fit to serological data from both cases and vaccinees (Figures 3A-C) were less likely to misclassify vaccinees as seroincident while maintaining sensitivity to detect recent infections.

**Figure 3:**
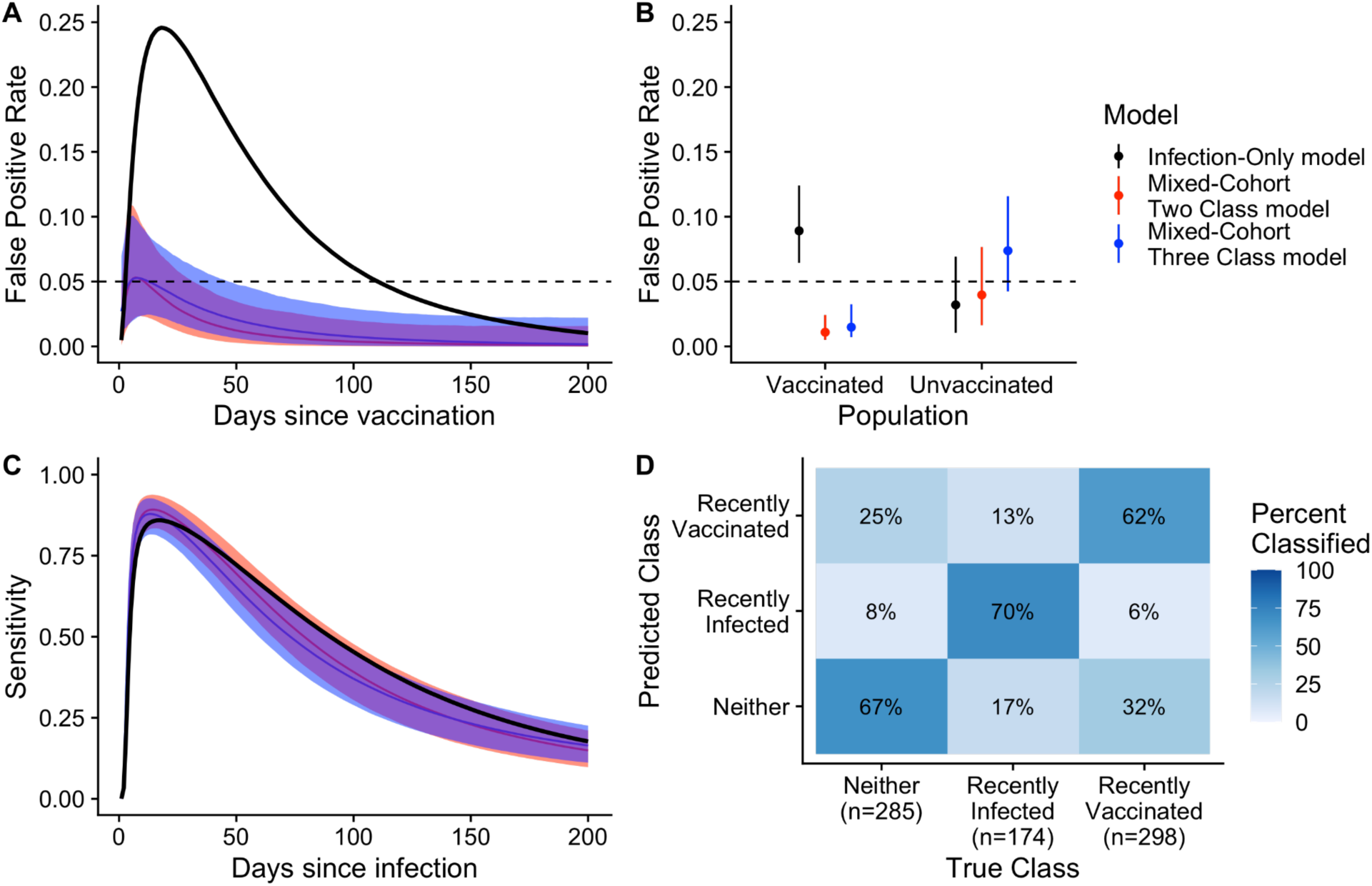
Comparison of performance of random forest models when serological data from vaccinees are included in the training set. Individuals were considered recently infected or vaccinated if exposed in the last 200 days. (A & B) Grey dashed line indicates the nominal false positivity rate of 5%. (A & C) Solid lines show the median value while shaded areas indicate the 95% credible interval. (D) Confusion matrix indicates the proportion of samples correctly classified from the new three-class model (*Mixed-Cohort Three Class Model*). Aside from the estimates for the false positivity rate among the vaccinated population for the *Case-only Model*, all other parameters were estimated through leave-one-individual-out cross-validation.

For both *Mixed-Cohort* models, the estimated false positivity rate among vaccinees was consistently below 5% between 0 and 200 days post vaccination (Figure 3A). Among vaccinees, the estimated false positivity rate (averaged over the 200-day period after vaccination) was much higher for the *Case-Only Model* (9%) than both *Mixed-Cohort* models (1-2%; Figure 3B). Among unvaccinated individuals, the estimated false positivity rate was only slightly lower for the *Case-Only Model* (3%) compared to the *Mixed-Cohort* models (4% and 6%) (Figure 3B). All three models had similar average sensitivity to detect individuals infected in the last 200 days (range: 45-55%) and similar patterns of time-varying sensitivity, peaking within the first 30 days after infection (Figure 3C). The addition of anti-O139 OSP and anti-TcpA antibody measurements, as well as IgA and IgM measurements of all antibodies, led to only marginal improvement in performance (Figure S4).

In some situations, it may be useful to not only distinguish between recently infected and not recently infected people, but also to identify recently vaccinated people from their serologic signatures. We investigated the performance of the *Mixed-Cohort Three Class Model* to further distinguish recently vaccinated (<200 days) individuals from those without a recent infection. The model correctly classified most samples, including 70% (121/174) from recently infected, 63% (188/298) from recently vaccinated and 68% (194/298) from individuals neither infected nor vaccinated recently (Figure 3D). Samples from both recently infected individuals (30/174, 17%) and recently vaccinated individuals (95/298, 32%) were more often misclassified as neither.

### Multiple adjustment strategies address bias in seroincidence estimates in partially-vaccinated populations

We simulated serological surveys conducted shortly after vaccination campaigns to assess the extent of bias in seroincidence estimates attributable to vaccination. Using the *Infection-Only Model* without accounting for increased misclassification among vaccine recipients, the magnitude of bias in seroincidence estimates varied with both the timing of the survey and the vaccination coverage. When the “true” 200-day incidence of infection was set at 10 infections per 100 individuals, serosurveys conducted 21 days post-campaign produced estimates with higher bias (i.e., the difference between estimated seroincidence and the true incidence of infection) as vaccination coverage rose. Specifically, the mean bias was 103% (range: 32– 185%) at 25% vaccination coverage and approximately 313% (range: 185–401%) at 75% coverage (Figure 4). In contrast, estimates based on the *Infection-Only Model* from serosurveys conducted 120 days after the vaccination campaign showed no discernible bias.

**Figure 4.**
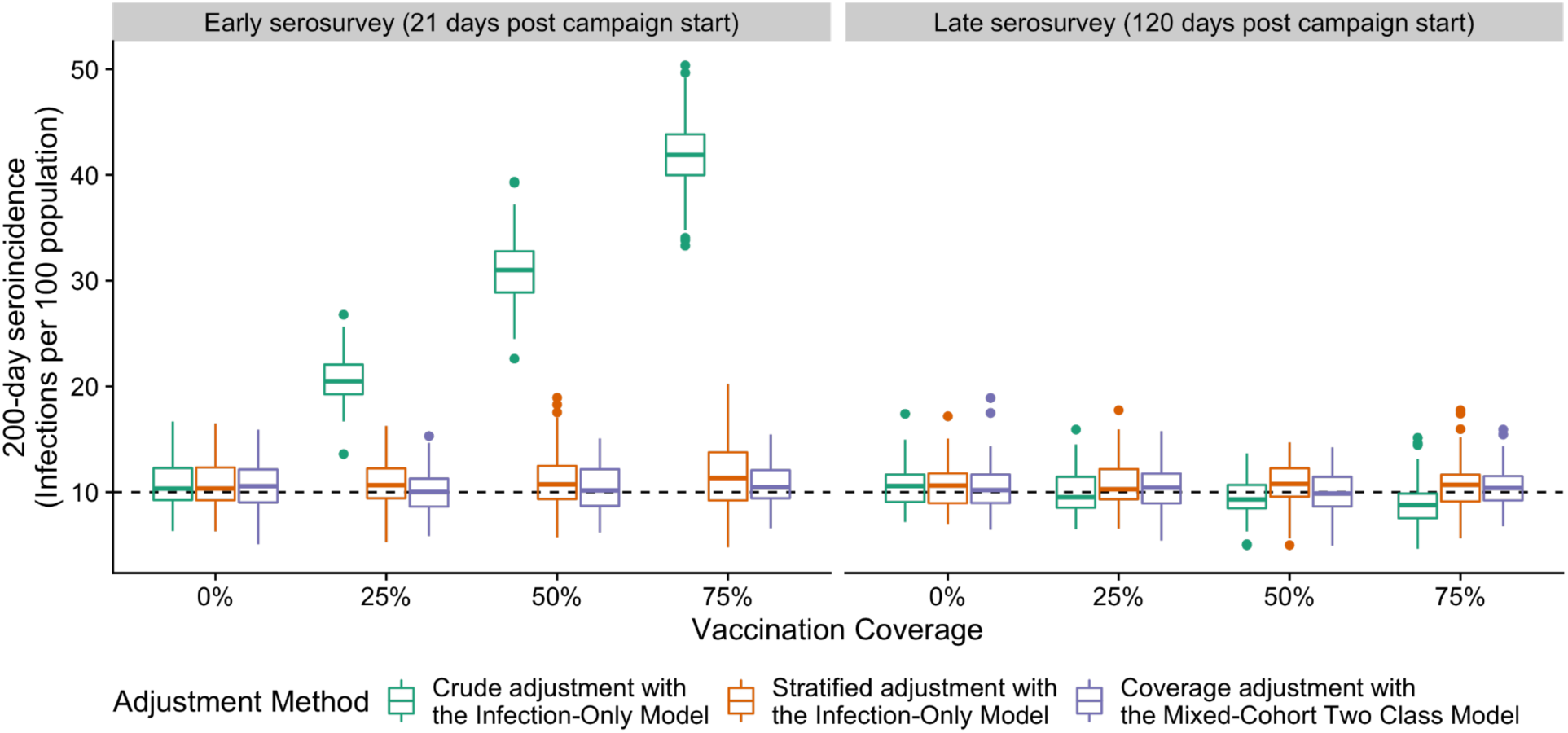
Simulated serological surveys in partially vaccinated settings using multiple strategies to calculate seroincidence 21 days (early) and 120 days (late) post vaccination campaign. Boxplots show the distribution of 100 estimates of seroincidence using different strategies at various levels of vaccine coverage (x-axis). Each simulated survey had 1000 persons. The true incidence was 10 infections per 100 in 200 days as indicated by the dashed line.

We found that two strategies (Table 3) mitigated bias in seroincidence estimates when surveys are conducted shortly after the vaccination campaigns: (1) use of the *Infection-Only* model adjusting for vaccine-status-specific performance and (2) use of the *Mixed-Cohort Two Class Model* with adjustment for model performance where the expected false-positive rate is informed by a vaccination coverage estimate. For example, at 50% vaccination coverage, adjusting for misclassification based on individual-vaccination status produced an average estimate of 11% (range: 4-21%), and using the *Mixed-Cohort Two Class Model* with an accurate estimate of vaccine coverage yielded an average estimate of 10% (range: 5-16%).

**Table 3.** Description of strategies for serosurveys to address misclassification due to vaccination campaigns.

| Name | Data collected (Sample size) | Prediction Model Used | Statistical adjustment of predictions |
| --- | --- | --- | --- |
| Crude adjustment with the *Infection-Only Model* | Serological survey (n=1000) | *Infection-Only model* | 1. Incidence rate is adjusted by sensitivity and false positivity rate to estimate overall seroincidence |
| Stratified adjustment with the *Infection-Only Model* | Serological survey & questionnaire data (n=1000) | *Infection-Only model* | 1. Vaccination status specific incidence rates are adjusted by sensitivity and false positivity rate<br>2. Vaccination status specific seroincidence rates are combined to estimate overall seroincidence |
| Coverage adjustment with the *Mixed-Cohort Two Class Model* | Serological data (n=1000)<br>Vaccine coverage data (n=500) | *Mixed-Cohort Two Class Model* | 1. Coverage data informs the expected false positive rate in the population<br>2. Incidence rate is adjusted by sensitivity and false positivity rate to estimate overall seroincidence |

## Discussion

Despite similarities in the immune response between infection and vaccination, our work illustrates that estimating cholera seroincidence through serological surveillance is feasible in partially vaccinated populations. We found that vaccination stimulates only a subset of the antibody repertoire of that generated by infection and to a lower degree. Due to these different post-exposure kinetics, excess misclassification of recently vaccinated individuals as seroincident only occurs when samples are collected within three months of vaccination. Accurate estimation of seroincidence can be made with a new generation of models proposed in this paper, simply by waiting a few months before conducting a serosurvey, or through collecting and incorporating data on vaccination coverage, at the individual or population level.

Vaccination and infection lead to measurable differences in the antibody response, which can be harnessed for serological surveillance. Though anti-Ogawa OSP and anti-Inaba OSP antibodies were generated among both cohorts, vaccinees generally had lower boosts overall. CTB and TcpA IgG and IgA antibodies were only stimulated by infection, and their presence could be used to distinguish infection from vaccination though misclassification with this approach could occur with children, who may have higher levels of anti-CTB antibodies (due to ETEC infection), and are generally less likely to generate anti-TcpA antibodies than adults (13). Lastly, we found that few anti-O139 OSP antibodies were generated from vaccination (similar to previous findings (14)) and are unlikely to effectively differentiate vaccinees from infected individuals). Therefore, we do not expect vaccination with a monovalent version of OCV without an O139 strain, which was WHO-prequalified in mid-2024, to be any more challenging to differentiate from infection than current bivalent OCV (15).

Stimulation of OSP antibodies through vaccination sometimes led to overestimation of seroincidence, but we proposed multiple ways to address this in the context of cross-sectional serological surveys. First, we found that models trained to identify individuals infected in the last 120 days sometimes misclassified vaccinated individuals as recently infected, though this slightly elevated misclassification occurred only briefly (i.e. within 49 days). As the duration of most cholera outbreaks have been documented to be less than 120 days (16), estimating incidence in a cross-sectional survey shortly after an outbreak ends may have minimal bias using standard seroincidence models, especially if the vaccination campaign occurs early on in the outbreak. For situations where an infection window of 200 days is being considered, we found that misclassification can likely be addressed either through using new seroincidence models where vaccinated individuals have been used in the training sets and/or through ascertaining individual-level vaccination status (e.g. through a questionnaire). However, the adjustment we proposed assumes that all individuals are vaccinated within a short window of time and requires estimates of sensitivity and specificity for the classification models. When starting a cross-sectional serosurvey, models to identify recent infections would ideally be validated with longitudinal serum samples from cases and vaccinees from a similar setting and target population.

This study comes with several notable limitations. The differences in the serological data among cases and vaccinees in this study may be partially attributed to differences in the age distribution and history of cholera spread in the area shortly before sample collection. Also, our study population only contained individuals who were infected with *V. cholerae* O1 or received two doses of OCV, though none with both a known recent infection and vaccination. As cholera vaccination becomes more common, future serological studies will allow the capture of post-infection kinetics of individuals with a history of vaccination. Challenge studies, where exposure can be directly controlled, might also be well-suited for gathering serological data on individuals with different combinations of exposures. We did not investigate ETEC infection, which is prevalent in many locations where cholera is, stimulates cross-reactive antibodies that can bind to CTB (17), and may also cause excess misclassification by seroincidence models. We also did not investigate the dynamics after only single dose vaccination though previous work indicates limited additional immunogenicity from the second dose though decay rates could differ (18, 19). Lastly, though our simulation results indicate there are reasonable strategies to address excess misclassification due to vaccination, these simulations rely on several assumptions including that individuals both vaccinated and infected would have the same antibody dynamics as someone who was not vaccinated but infected.

Estimating the incidence of infection and/or disease is essential for identifying priority areas for cholera prevention, control measures, and the allocation of scarce resources like vaccines. Seroincidence estimation using cross-sectional serosurveys offers a valuable method to assess infection incidence, especially in settings where clinical surveillance data may be unreliable. Our study highlights several promising strategies to mitigate the overestimation of cholera seroincidence in partially vaccinated populations. While additional data across diverse populations are needed to evaluate the generalizability of our findings, these results suggest that the proposed methods are feasible and practical in the increasingly vaccinated landscape of cholera-affected regions.

## Methods

### Study Population

In this study we analyzed data from three cohorts: confirmed cholera cases and their household contacts enrolled in Bangladesh, volunteers vaccinated with Shanchol enrolled in Bangladesh and Haiti (i.e. vaccinees).

In Bangladesh, as described previously, consenting patients greater than 1 years old hospitalized at the International Centre for Diarrhoeal Disease Research, Bangladesh (icddr,b) Dhaka hospital with culture confirmed *V. cholerae* O1 were enrolled between 2006 and 2018 (20, 21). We utilized data from a previous analysis where we selected 51 confirmed cases (2 - 1080 days post symptom onset) and uninfected contacts enrolled in Bangladesh (309 serum samples in total) (12), ensuring representation from both children and adults.

A non-inferiority trial comparing two killed, whole cell cholera vaccines (Shanchol and Cholvax, Incepta [a locally produced OCV in Bangladesh])) was conducted in Mirpur, Bangladesh between April 2016 and April 2017 (22). We selected a convenience sample of vaccinees that received Shanchol, including both children and adults, to test for this study. Serum samples were collected up to 42 days after the first dose of vaccination through 6 study visits.

In Haiti, serum samples were collected from healthy volunteers enrolled from the outpatient department at Saint Nicholas Hospital in St. Marc, Haiti, an urban center in the Artibonite Department (23). Individuals were excluded if previously given OCV, pregnant, or reported to have had active gastrointestinal disorder within 7 days prior to enrollment. All individuals received two doses of kOCV spaced 14 days apart. All vaccinees were ≥18 years old enrolled in 2015 (May) and 2016 (January, March and April). Samples were collected around 0, 7, 21, 44, 90-, 180-, 270-, and 365-days post first dose. While cholera cases were reported regularly in Haiti after being introduced in 2010, cases steadily decreased from 2011 through the time of these studies (24). We had access to samples of 73 vaccinees followed up to 360 days. After limiting selection to 67 individuals (92%) with at least 3 samples, we randomly selected 36 vaccinees.

All samples from patients, contacts and vaccinees were collected in fully IRB-approved studies following informed consent. Samples from children were collected following informed consent of the parent/guardian. These studies were approved by the Research Review Committee and Ethical Review Committee of the icddr,b, the IRB of the Massachusetts General Hospital (2013P002604) and the Haitian National Bioethics Committee (Ref:1415-81).

### Serological testing and data processing

Based on a review of the published literature on immune responses to *V. cholerae* infection (12, 25–27), we focused our analysis on five cholera-related antigens using a multiplex bead assay. These included O1 serogroup Ogawa serotype O-specific polysaccharide (OSP-BSA, part of the LPS), O1 serogroup Inaba serotype OSP, Cholera Toxin B-subunit (CTB), Toxin co-regulated pilus subunit A (TcpA), and O139 OSP (*V. cholerae* O139 serogroup is no longer a cause of significant numbers of human *V. cholerae* infections yet is included in the most commonly used bivalent OCVs). While not analyzed in this study, plates also included beads for cholera toxin holotoxin (CTH), *V. cholerae* cytolysin (VCC) (also known as hemolysin A), *V. cholerae* sialidase, heat-labile enterotoxin subunit B (LTB), and heat-labile holo-enterotoxin (LTH) (expressed during ETEC infection), and influenza hemaglutinin 1 (as a control antigen). All antigens were conjugated to Luminex magnetic beads as previously described (12).

All plates included a dilution series (from pooled convalescent sera of culture confirmed *V. cholerae* O1 cases in Bangladesh) and control wells, all of which were run in triplicate. Following the testing protocol, serum, beads, and secondary antibodies binding to IgG, IgA, and IgM were added to each well. Given the limited amount of O139 serogroup OSP available, we were unable to test all samples for O139 OSP IgA and IgM. Samples were run on a Luminex Flexmap 3D machine at Massachusetts General Hospital. Bead counts and median fluorescence intensity (MFI) values were exported from the Exponent software program. Plates were retested when over half of the positive control dilutions had >=5 antigens (excluding O139 OSP as they were not on all plates) with a coefficient of variation (calculated from triplicate MFI measurements) greater than 20%.

For the analysis, any measurements with a bead count of less than 30 were excluded (<0.5% of data points). MFI values were averaged across replicate wells. We standardized MFI values from the assay to help adjust for interplate variability by calculating the relative antibody unity (RAU) (28). For each plate, we fit a log-logistic model to the dilution series and used the median parameter estimates to predict the RAU for each sample (29, 30). For samples with a predicted RAU outside the range of 10^5^ and 10^2^, the RAU was set at the threshold value.

The complete laboratory protocol for the MBA assay is available at https://doi.org/10.17504/protocols.io.3byl4b1x8vo5/v1.

### Statistical Analyses

We compared the antibody kinetics of vaccinated volunteers and confirmed cholera cases by describing the average baseline, the geometric mean RAU ratio of the peak versus baseline of antibody measurements, and the proportion of individuals with a RAU ratio greater than 2-fold. We used multidimensional scaling (31, 32) to visualize the immunologic profiles (in two dimensions) of samples from individuals infected or vaccinated within the last 200 days, including all markers previously mentioned.

We trained random forest classification models to identify if a sample was from a recently infected individual based on antibody measurements. We define samples classified as recently infected as seroincident. We explored several different periods for defining a recent infection (<45 days, <120 days, <200 days & <300 days). The models relied on measurements of three IgG markers (anti-Ogawa OSP, anti-Inaba OSP, and anti-CTB) shown to rise after infection, which current seroincidence models depend on (12). In supplemental analyses, we also explored model performance where we included IgG, IgM, and IgA markers (anti-Ogawa OSP, anti-Inaba OSP, anti-CTB, and anti-TcpA) as well as anti-O139 OSP IgG measurements in the models.

Each random forest model contained 1000 classification trees, each of which “votes” for a sample as recently infected or not. Instead of choosing the class (i.e., seroincident or not) identified by the majority of votes, we established cut-offs for a nominal 5% false positivity rate among unvaccinated individuals. Specifically, we randomly selected 50% of the samples from unvaccinated individuals, trained a random forest model to predict serostatus, and selected the lowest cutoff (e.g., proportion of votes that need to predict positive to classify a result as positive) that led to no more than 5% of unvaccinated true-negative individuals in the held-out set to be misclassified as seroincident. We repeated this cross-validation procedure for each model 100 times and took the median of the cut-off values for each.

We fit three different types of random forest models: *Case-Only Model, Mixed-Cohort Two Class Model, and Mixed-Cohort Three Class Model* (Table 2). The *Case-Only Model* (used in Jones et al. (12)) is based on training data only from cholera cases and their household contacts in Bangladesh. The *Mixed-Cohort Two Class Model* is also trained on these data as well as vaccinee data which are not considered recently infected. The *Mixed-Cohort Three Class Model* is also trained on case, household contact, and vaccinee data, but samples are classified as recently infected (<200 days), recently vaccinated (<200 days), or neither. When the *Mixed-Cohort Three Class Model* classifies a sample, it simply selects the class with the highest number of votes among the three classes.

We assessed accuracy of random forest models using leave-one-individual-out cross validation (LOOCV) to calculate the time-varying false positivity rate among vaccinees, time-varying sensitivity among cases, and the false positivity rate among unvaccinated individuals. For time-varying false positivity and time-varying sensitivity models, we assumed that the logit (proportion seroincident) was a cubic function of (natural log-transformed) days since infection with a random-intercept for each individual. For the false positivity rate among unvaccinated individuals, we assumed that the logit (positivity) was only a random-intercept (i.e., per person) model. We report 95% credible intervals (CI).

To evaluate strategies for mitigating bias in seroincidence estimates, we simulated serosurveys to estimate 200-day seroincidence at both 21 days (early) and 120 days (late) after vaccination campaigns (Table 3). We evaluated three strategies: Crude adjustment with the *Infection-Only Model* (i.e., no adjustment taking into account vaccination status), stratified adjustment with the *Infection-Only Model* (i.e., adjust estimates for differential model performance by vaccination status of the individual), and coverage adjustment with *Mixed-Cohort Two Class Model* (i.e., predict who is recently infected with the *Mixed-Cohort Two Class Model*). We ran 100 simulations per scenario.

In each simulated serosurvey, 1000 participants were randomly selected from an area with a constant infection incidence of 10%. Vaccination coverage was set at variable levels (0%, 25%, 50%, and 75%). Once vaccination status and days since last infection were randomly drawn for each person, we used individual-level estimates of sensitivity and specificity to simulate their seroincident status for the random forest model used. Individuals who were both recently infected and vaccinated were assumed to have the immunological profile of those who were recently infected but not vaccinated. We then calculated estimates of seroincidence after adjusting for test misclassification.

## Data Availability

All data produced in the present study are available upon reasonable request to the authors

## Data and code availability

Data and code used to conduct statistical analyses are available at https://github.com/HopkinsIDD/cholera-vaccine-serosurvey.

## Acknowledgments

We thank Rachel Mills for her help with laboratory analyses. We thank the Infectious Disease Dynamics Group at Johns Hopkins University for insight on the statistical analysis. We are grateful to Dr. Slavomír Bystrický, Institute of Chemistry, Slovak Academy of Sciences, Bratislava, Slovak Republic, and Paul Kováč and Peng Xu from the National Institutes of Health for the provision of OSP-BSA reagents. We acknowledge the study participants and families who consented to enroll into this study.

---

**Table S1.** Timing of sample collection for each cohort.

| Exposure Type | Population | Days since exposure | Individuals | Samples (%) |
| --- | --- | --- | --- | --- |
| Vaccination with Shanchol | Bangladeshi volunteer | 0 | 43 | 43 (100) |
| Vaccination with Shanchol | Bangladeshi volunteer | 3 | 43 | 40 (93) |
| Vaccination with Shanchol | Bangladeshi volunteer | 14 | 43 | 40 (93) |
| Vaccination with Shanchol | Bangladeshi volunteer | 17 | 43 | 40 (93) |
| Vaccination with Shanchol | Bangladeshi volunteer | 28 | 43 | 37 (86) |
| Vaccination with Shanchol | Bangladeshi volunteer | 42 | 43 | 36 (84) |
| Vaccination with Shanchol | Haitian volunteer | 0 | 36 | 36 (100) |
| Vaccination with Shanchol | Haitian volunteer | 7 | 36 | 36 (100) |
| Vaccination with Shanchol | Haitian volunteer | 21 | 36 | 35 (97) |
| Vaccination with Shanchol | Haitian volunteer | 44 | 36 | 22 (61) |
| Vaccination with Shanchol | Haitian volunteer | 90 | 36 | 34 (94) |
| Vaccination with Shanchol | Haitian volunteer | 180 | 36 | 18 (50) |
| Vaccination with Shanchol | Haitian volunteer | 220 | 36 | 8 (22) |
| Vaccination with Shanchol | Haitian volunteer | 360 | 36 | 23 (64) |
| Natural infection with *V. cholerae* O1 | Bangladeshi case-patient | 2 | 48 | 48 (100) |
| Natural infection with *V. cholerae* O1 | Bangladeshi case-patient | 7 | 48 | 46 (96) |
| Natural infection with *V. cholerae* O1 | Bangladeshi case-patient | 30 | 48 | 46 (96) |
| Natural infection with *V. cholerae* O1 | Bangladeshi case-patient | 90 | 48 | 42 (88) |
| Natural infection with *V. cholerae* O1 | Bangladeshi case-patient | 180 | 48 | 40 (83) |
| Natural infection with *V. cholerae* O1 | Bangladeshi case-patient | 270 | 48 | 12 (25) |
| Natural infection with *V. cholerae* O1 | Bangladeshi case-patient | 360 | 48 | 14 (29) |
| Natural infection with *V. cholerae* O1 | Bangladeshi case-patient | 540 | 48 | 25 (52) |
| Natural infection with *V. cholerae* O1 | Bangladeshi case-patient | 720 | 48 | 1 (2) |
| Natural infection with *V. cholerae* O1 | Bangladeshi case-patient | 900 | 48 | 25 (52) |
| Natural infection with *V. cholerae* O1 | Bangladeshi case-patient | 1,080 | 48 | 1 (2) |
| Household member of case-patient infected with *V. cholerae* O1 | Bangladeshi volunteer | 2 | 3 | 3 (100) |
| Household member of case-patient infected with *V. cholerae* O1 | Bangladeshi volunteer | 7 | 3 | 3 (100) |
| Household member of case-patient infected with *V. cholerae* O1 | Bangladeshi volunteer | 30 | 3 | 3 (100) |

**Figure S1.**
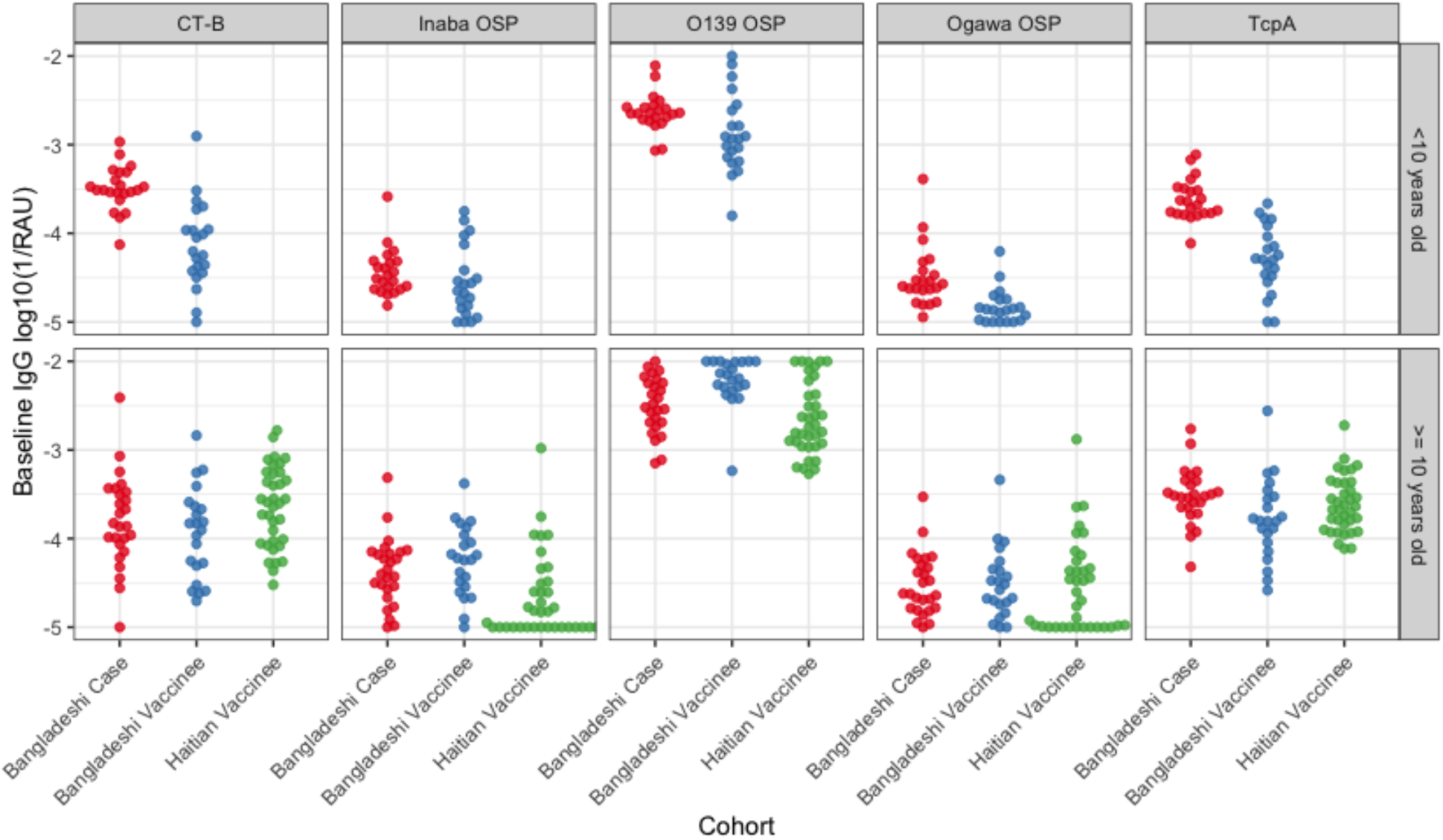
Baseline comparison of IgG markers.

**Table S2.** Geometric mean fold-rise in RAU by marker among cases and vaccines.

| Antigen | Isotype | Case | Vaccinee |
| --- | --- | --- | --- |
| CT-B | IgA | 23.3 | 1.2 |
| CT-B | IgG | 23.9 | 1.5 |
| CT-B | IgM | 1.5 | 1.1 |
| Inaba OSP | IgA | 15.1 | 6.6 |
| Inaba OSP | IgG | 12.5 | 6.1 |
| Inaba OSP | IgM | 13.2 | 4.4 |
| O139 OSP | IgA | 2.1 | 2.0 |
| O139 OSP | IgG | 1.7 | 1.6 |
| O139 OSP | IgM | 1.5 | 1.4 |
| Ogawa OSP | IgA | 24.5 | 8.2 |
| Ogawa OSP | IgG | 33.4 | 9.7 |
| Ogawa OSP | IgM | 44.9 | 6.7 |
| TcpA | IgA | 3.4 | 1.3 |
| TcpA | IgG | 4.1 | 1.7 |
| TcpA | IgM | 1.8 | 1.2 |

**Table S3.**
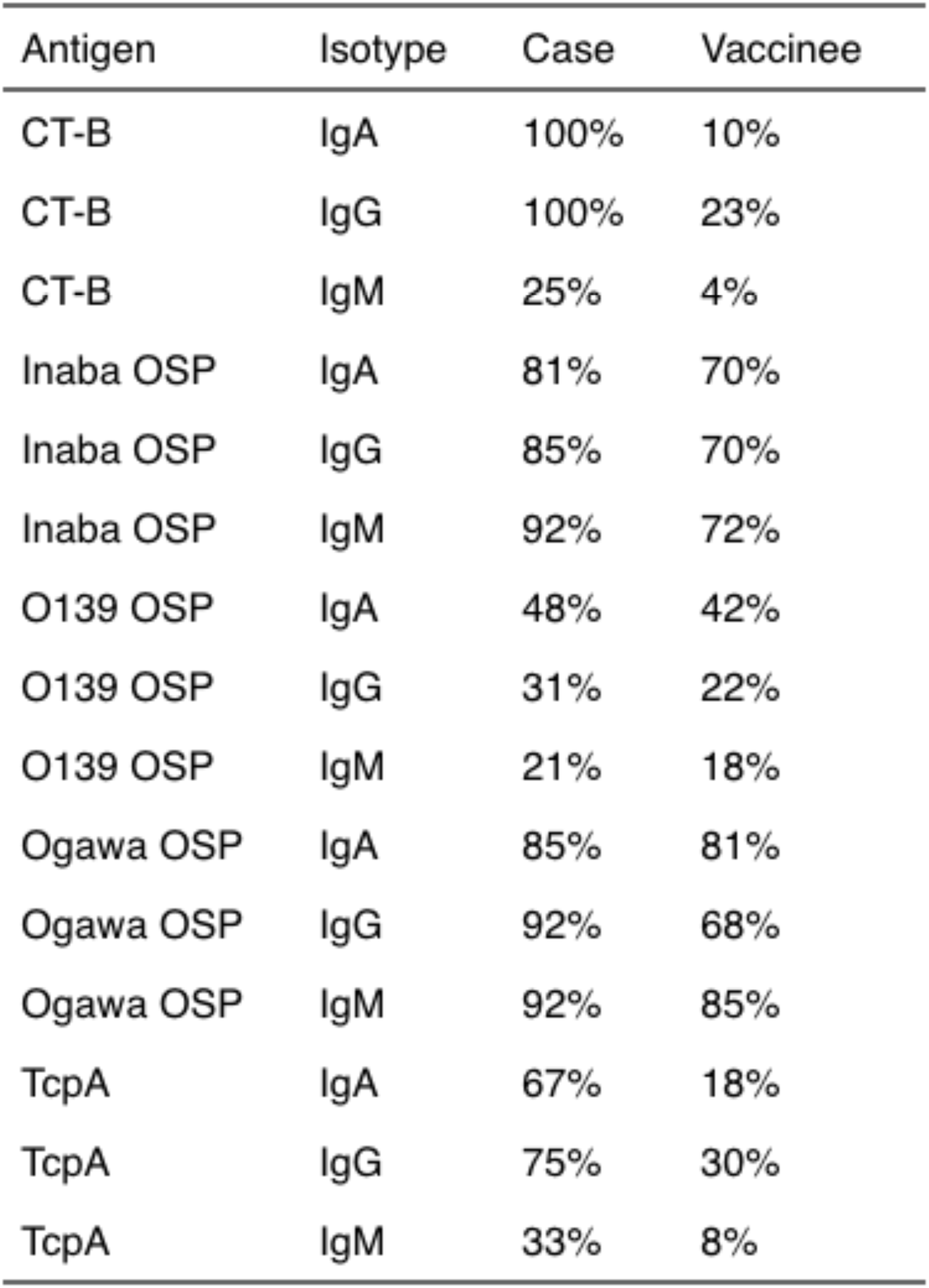
Proportion of individuals with a two fold-rise in RAU by marker among cases and vaccines.

| Antigen | Isotype | Case | Vaccinee |
| --- | --- | --- | --- |
| CT-B | IgA | 100% | 10% |
| CT-B | IgG | 100% | 23% |
| CT-B | IgM | 25% | 4% |
| Inaba OSP | IgA | 81% | 70% |
| Inaba OSP | IgG | 85% | 70% |
| Inaba OSP | IgM | 92% | 72% |
| O139 OSP | IgA | 48% | 42% |
| O139 OSP | IgG | 31% | 22% |
| O139 OSP | IgM | 21% | 18% |
| Ogawa OSP | IgA | 85% | 81% |
| Ogawa OSP | IgG | 92% | 68% |
| Ogawa OSP | IgM | 92% | 85% |
| TcpA | IgA | 67% | 18% |
| TcpA | IgG | 75% | 30% |
| TcpA | IgM | 33% | 8% |

**Figure S2:**
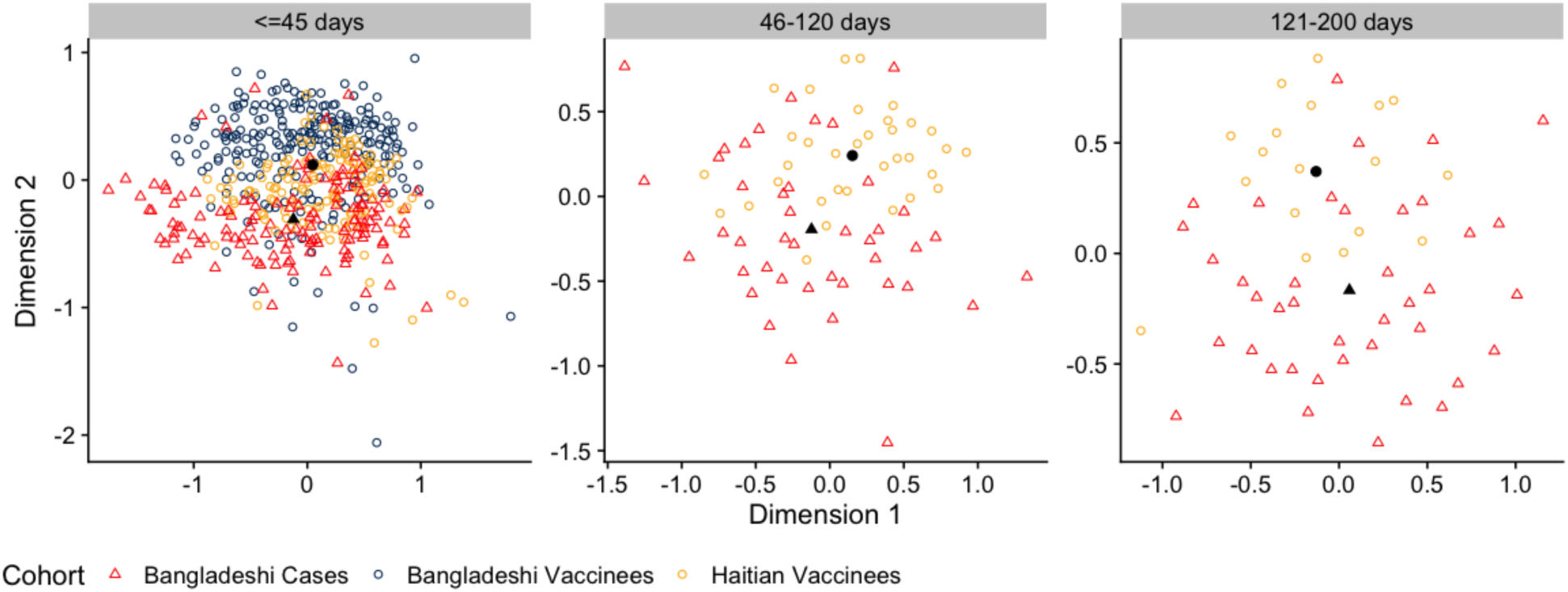
Multidimensional scaling analysis of all serological data collected. Each panel includes the two dimensions calculated from data collected within the specified time window. Black points represent the location of the centroids for the two new dimensions of cases (circle) and vaccinees (triangle).

**Figure S3:**
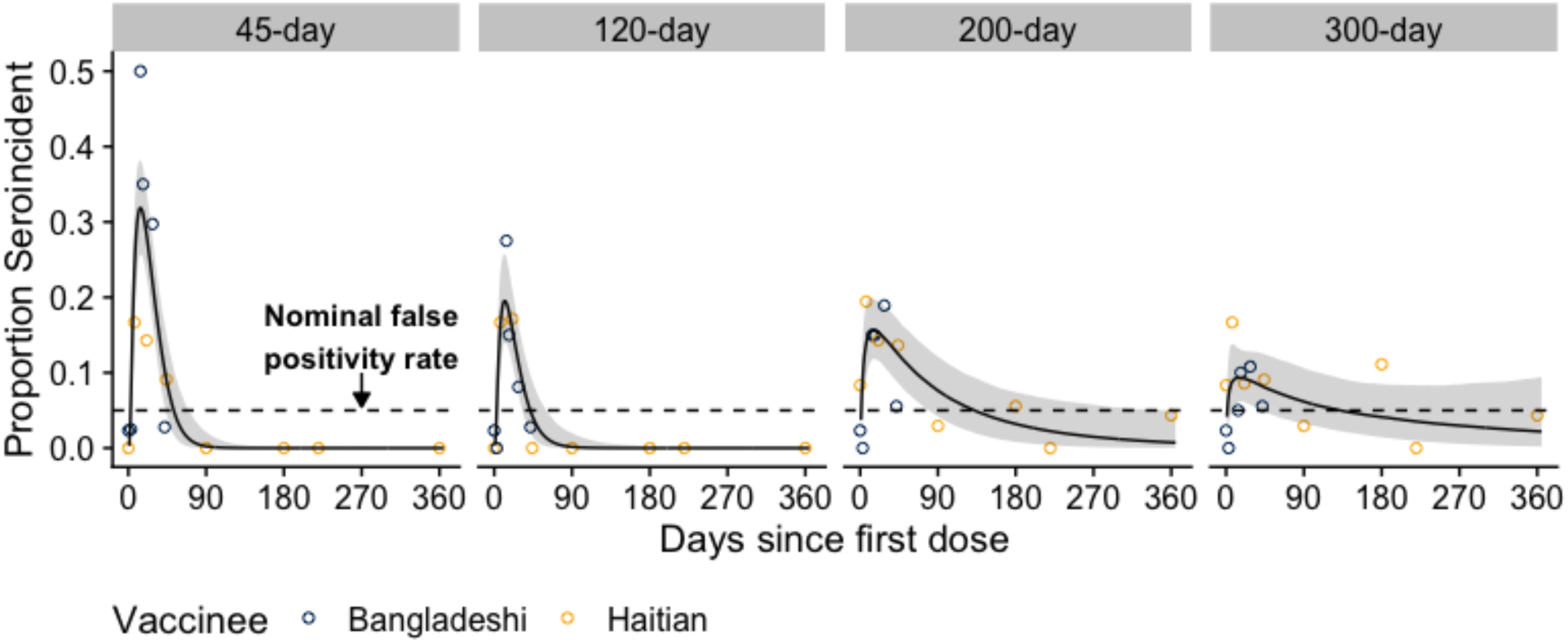
Misclassification of vaccinees as seroincident by previous seroincidence models with additional markers using 45, 120, 200, and 300 day infection windows. The model used (i.e., the Case Only Model) to classify vaccinees as seroincident or not was a previously published random forest model trained on anti-CTB, anti-Ogawa OSP, anti-Inaba OSP, anti-TcpA antibodies (IgG, IgM, and IgA) as well as anti-O139 OSP IgG from Bangladeshi confirmed cases and uninfected household contacts. The proportion of Bangladeshi (dark blue) and Haitian vaccinees (gold) classified as seroincident are shown as dots. The overall proportion seroincident was modeled with a cubic spline (black line and grey ribbon) using data from both cohorts of vaccinees. Black dashed line indicates the nominal false positivity rate of 5%.

**Figure S4:**
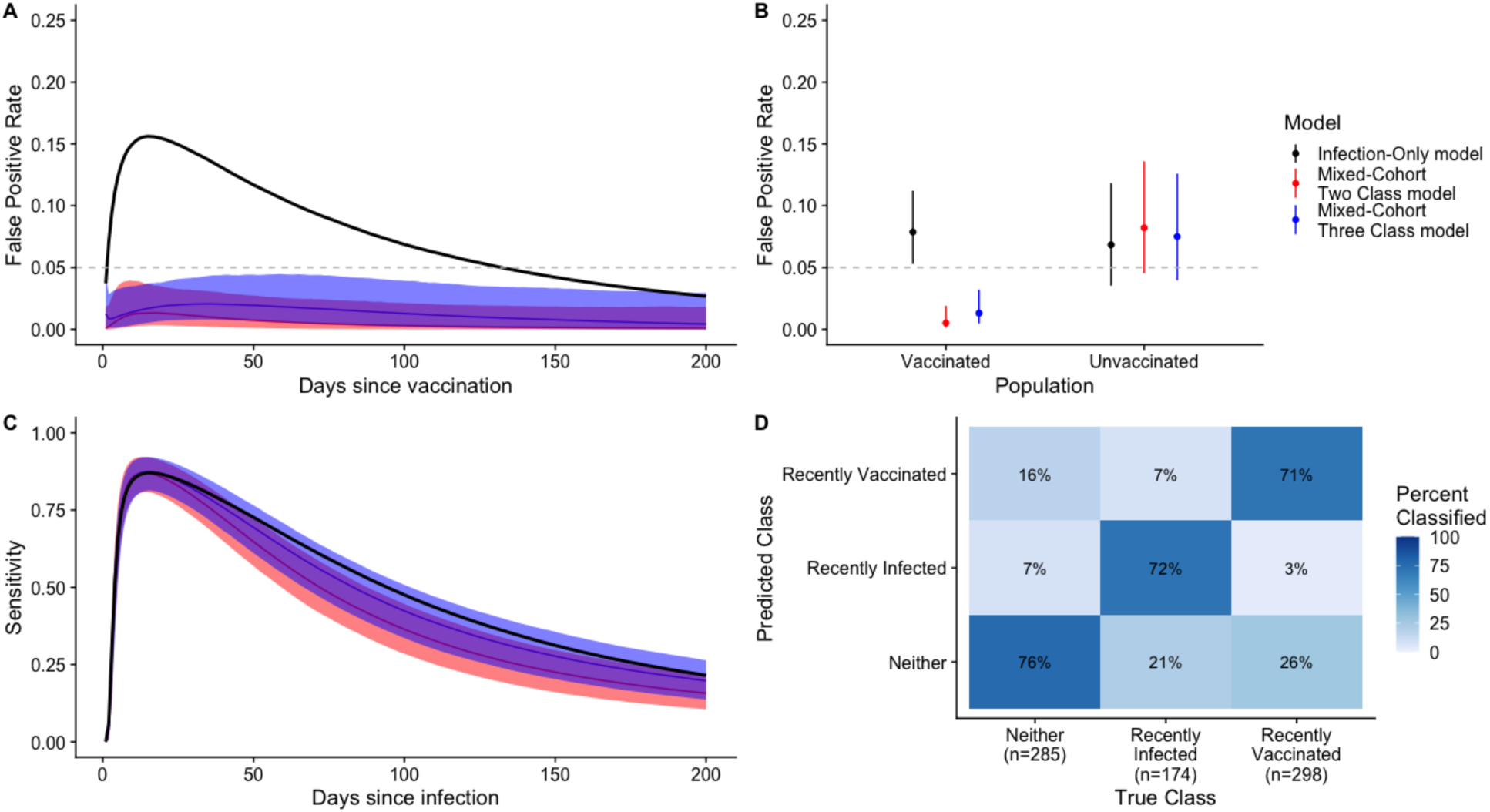
Comparison of performance of random forest models when all IgG, IgM, and IgA serological data from vaccinees are included in the training set. Individuals were considered recently infected or vaccinated if exposed in the last 200 days. (A & B) Grey dashed line indicates the expected/nominal false positivity rate of 5%. (A & C) Solid lines show the median value while shaded areas indicate the 95% credible interval. (D) Confusion matrix indicates the proportion of samples correctly classified from the new three-class model (*Mixed-Cohort Three Class Model*). Aside from the estimates for the false positivity rate among the vaccinated population for the *Case-only Model*, all other parameters were estimated through leave-one-individual-out cross-validation.

